# Characterization and Racial Stratification of Social Determinants of Health for Individuals with Type 2 Diabetes as Recorded in Electronic Health Records: Implications for Artificial Intelligence Development

**DOI:** 10.1101/2024.10.07.24315048

**Authors:** Polina V. Kukhareva, Matthew J. O’Brien, Daniel C Malone, Kensaku Kawamoto, Ramkiran Gouripeddi, Deepika Reddy, Mingyuan Zhang, Vikrant G. Deshmuch, Julio C. Facelli

**Author notes:** **Corresponding author:** Polina Kukhareva, PhD, MPH, Assistant Professor, Department of Biomedical Informatics, University of Utah, Salt Lake City, UT, USA, 421 Wakara Way, Suite 108, Salt Lake City, UT 84108. **Funding Statement** This research received no specific grant from any funding agency in the public, commercial, or not-for-profit sectors. JCF and RG were partially funded by the National Center for Advancing Translational Sciences of the National Institutes of Health under Award Number UM1TR004409. MJO was partially funded by the National Institute of Diabetes and Digestive and Kidney Diseases under Award Number P30DK092949. **Contributorship Statement** PVK, MJO, KK and JCF drafted the manuscript. PVK takes responsibility for the manuscript’s content, including the data and analysis. Each author contributed substantially to the drafting or substantial revision of the paper. All authors contributed significantly to the study design, data interpretation, and manuscript writing. All authors also approved the paper for submission and agreed both to be personally accountable for the author’s own contributions and to ensure that questions related to the accuracy or integrity of any part of the work are appropriately investigated, resolved, and documented in the literature. PVK had full access to data. PVK conducted the statistical analyses.

## Abstract

**Background:** Accurate documentation of social determinants of health (SDoH) in electronic health records (EHRs) is critical for developing equitable AI models for diabetes management. This study investigates SDoH data in a cross-institutional EHR database.

**Methods:** We analyzed neighborhood-level (i.e., social vulnerability index [SVI], Rural-Urban Community Area [RUCA]) and individual-level SDoH (e.g., preferred language, marital status, tobacco, alcohol, and substance use) within the Epic Cosmos database, focusing on adults diagnosed with T2D (E11.*) who had encounters between 2021 and 2023. We measured data completeness (i.e., the proportion of individuals who have a non-missing value) and the prevalence of non-canonical values (e.g., preference for language other than English) for each available SDoH variable.

**Findings:** The study included 12,696,680 individuals with T2D. SVI, RUCA and preferred language were available for all individuals, while marital status, and smoking data were available for over 90%. However, financial needs, interpersonal violence, social activity, and physical activity were present in EHRs for 7.6%-24.6% of the population depending on race/ethnicity. Minority groups experienced lower data completeness and higher burden of non-canonical values compared to White individuals.

**Interpretation:** Neighborhood-level and some individual-level SDoH have potential for use in AI development and evaluation. Other SDoH data cannot be used without additional analysis to address high amounts of missing data. Significant disparities in completeness exist across racial/ethnic groups. Addressing these data gaps may require government and payer mandates, standardized SDoH screening tools, and personnel training.

**Highlights:**

1. This study examined social determinants of health (SDoH) data for adults with type 2 diabetes in a cross-institutional electronic health record (EHR) database to support equitable AI model development.
2. Neighborhood-level SDoH data and some individual-level SDoH data (individual-level SDoH (i.e., race/ethnicity, preferred language, marital status) were highly complete.
3. Disparities in SDoH data completeness by race/ethnicity underscore the need for standardized SDoH documentation.

## Background

Type 2 diabetes (T2D) is a chronic metabolic condition that affected 38.4 million U.S. adults in 2021 and is expected to affect 60.6 million by 2060.^1,2^ To address this public health crisis, the use of artificial intelligence (AI) for managing T2D in clinical settings is on the rise.^3^ For example, several models were recently developed to provide personalized T2D medication recommendations.^4^ Electronic health records (EHRs), with almost universal adoption and growing data interoperability, serve as an essential source of data for clinical AI algorithms.^4^

While use of AI models based on EHR data is an promising and exciting development,^4^ there is a growing concern that AI models may exacerbate existing healthcare disparities by underperforming in populations that are already vulnerable.^5–10^ Biases inherent in the training data, such as lack of representation and informativeness for certain demographic groups, can lead to proposing suboptimal treatment plans.^11^ Despite these concerns, there has been relatively little effort to systematically identify and mitigate bias in clinical ML models,^12^ making it crucial to address these issues to ensure equitable healthcare for all populations.

The prevalence of T2D is nearly two times higher in racial and ethnic non-White groups than among White Americans, highlighting health inequities that may be shaped in part by the social determinants of health (SDoH), the conditions in which people are born, grow, live, work, and age.^13–16^ These disparities make it especially important to ensure that the emerging AI-algorithms for clinical management of T2D are fair. AI fairness refers to ensuring that AI models perform equitably across different demographic groups. High completeness of SDoH variables is essential to evaluate fairness of emerging AI algorithms.

EHRs could potentially become a primary source of SDoH data for validation of AI algorithms.^17^ Several healthcare stakeholders, including CMS and the Joint Commission, mandate the collection of certain SDoH data, and many health systems are voluntarily striving to collect this information. Epic, which is used by approximately half of all ambulatory medical providers, is prioritizing the collection of SDoH data,^18^ and there are many ongoing efforts to standardize the collection of SDoH data in EHRs using validated questionnaires.^19–21^ Additionally, data interoperability standards are starting to be used to store and exchange SDoH data in a consistent way.^22^ However, while early reports indicate that the utility of such SDoH data is limited by low completeness,^20^ to the authors knowledge, no comprehensive studies stratified by race/ethnicity in multi-institutional data repositories have been reported for individuals with T2D.

This study aimed to address whether SDoH variables are sufficiently present in EHRs to enable accurate AI fairness evaluation in a very large multi-institutional EHR data repository, Epic Cosmos.^23^

## Methods

### Setting

Data used in this study came from Epic Cosmos,^24^ a community collaboration of health systems representing over 251 million individual records from over 1,400 hospitals and 32,500 clinics. Epic Cosmos includes aggregated data from a substantial percentage of the U.S. population. Data extraction was completed on June 14, 2024.

### Study Population

We included adults (age 18 years or older at the data extraction date) who had an office visit, telemedicine encounter, surgery visit, or lab visit in 2021-2023 and had a T2D diagnosis code (E11.*) as an encounter diagnosis, billing final diagnosis or active in the problem list in 2015-2023. Combined race/ethnicity was aggregated into American Indian or Alaskan Native (AI/AN), Asian, Black or African American (Black), Hispanic or Latino (Hispanic), Native Hawaiian or Other Pacific Islander (NH/PI), and White. Non-white individuals were defined as those who reported race/ethnicity other than White or Caucasian. Persons identifying with more than one racial category were excluded from the analysis of individual-level SDoH.

### Neighborhood-level SDoH

Unlike individual-level SDoH variables, which are specific to the individual, neighborhood-level SDoH are based on the individual’s residence and may be less precise for an individual. We analyzed two neighborhood-level SDoH variables: Social Vulnerability Index (SVI) and Rural-Urban Community Area (RUCA) codes. The SVI helps public health officials and planners prepare communities for emergencies like severe weather, disease outbreaks, or chemical exposure. It is based on 16 U.S. census variables and includes four main categories: socioeconomic status, racial and ethnic minority status, household characteristics, and housing and transportation.^25^ SVI scores range from 0 (less vulnerable) to 1 (more vulnerable). RUCA codes categorize U.S. census tracts by urbanization, population density, and commuting patterns. In the EHR databases, neighborhood-level variables are imputed from the individual address, which is widely available.

### Individual-level SDoH

Individual-level SDoH can be documented in the EHR by clinical providers and medical support staff. Collection of SDoH data is standardized using integrated SDoH displays (**Figure 1**). We included all available SDoH variables that are currently available in Epic Cosmos. Available variables included preferred language, marital status, 12 variables related to the use of substances with potential for misuse and addiction (tobacco, alcohol, psychoactive drugs), 6 variables related to resource needs (financial resource strain, food scarcity, food worry, medical transportation needs, non-medical transportation needs, and housing instability), 4 variables related to interpersonal violence (emotional, fear, physical abuse, sexual abuse), 2 variables related to physical activity (days per week, minutes per session), 5 variables related to social connections (church, get together, meetings, membership, phone) and 1 attribute related to stress. For individuals who had multiple values recorded, we used the last value collected prior to the data extraction date.

**Figure 1.**
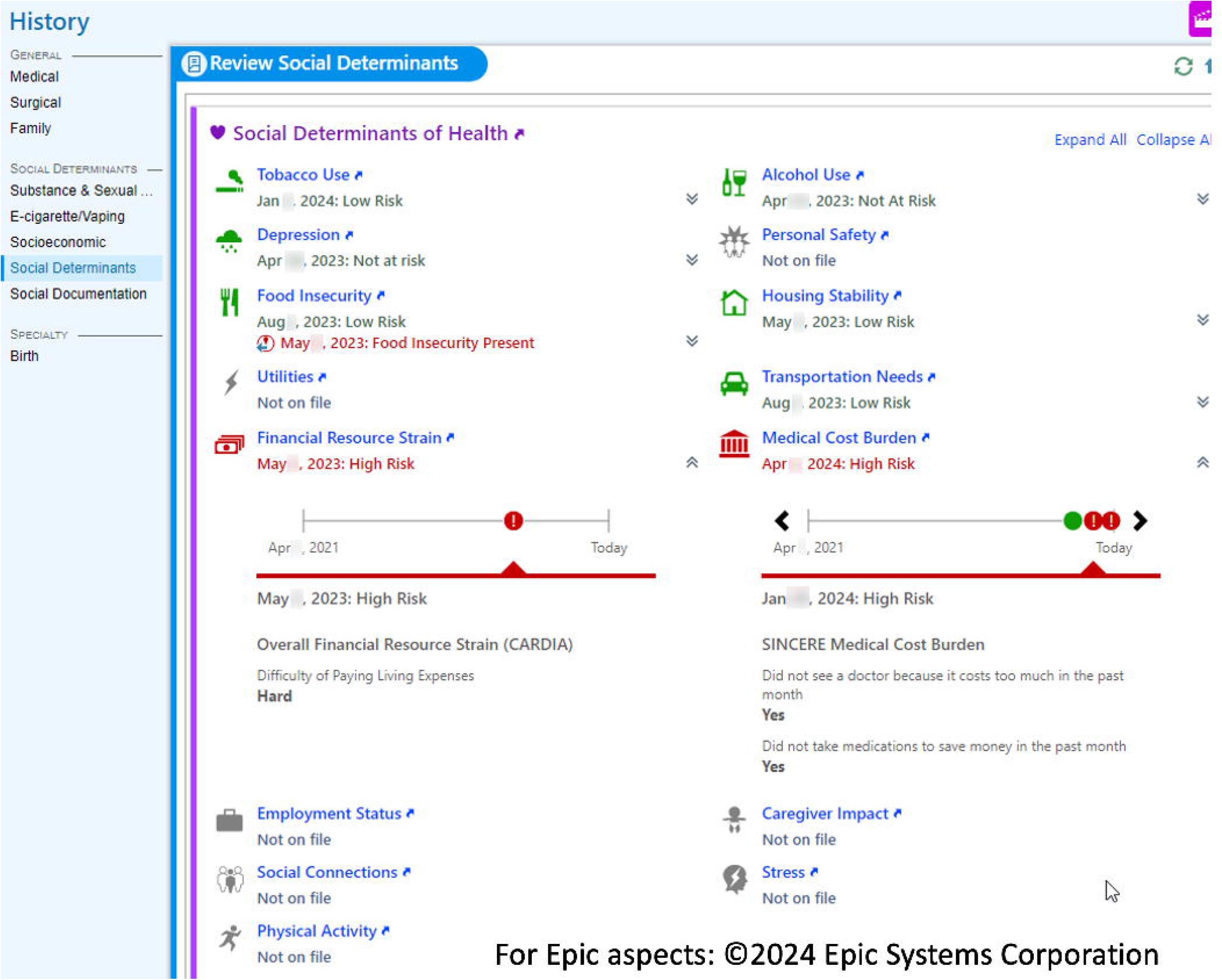
SDoH Data Captured and Displayed in the EHR.

### Study Measures

This study evaluated completeness and prevalence of non-canonical values for each SDoH variable. Completeness of an SDoH variable was defined as the proportion of individuals who have a non-missing value for this attribute. We also evaluated the completeness of detailed information for individuals who reported use of substances such as smoking history, alcohol use, and substance use. For example, we evaluated the completeness of detailed smoking history (pack-years and years smoked) for individuals who reported smoking. Prevalence of non-canonical answers referred to answers different from the canonical value. We defined canonical answers as those expected by the majority class (White), for instance English language:

- Preferred language: English
- Marital status: Married
- Tobacco Use, Smokeless; Tobacco Use, Smoking, Alcohol Use, and Substance Use: Never used
- Food Scarcity; Food Worry: Never true
- Transportation needs, housing instability, interpersonal violence: No issues
- Financial Resource Strain: Not hard at all
- Physical activity: More than 0 days and 0 minutes of activity
- Social activity: Any level of activity other than never
- Stress: Not at all stressed
- Cigarette smoking: Less than 20 pack-years and less than 1 pack per day
- Alcohol use: Less than 1 drink per day, less than 2 drinks per week, fewer than 3 standard drinks, and less than 1 binge per month
- Drug misuse: Once or less
- Substance abuse: Use of drugs other than the following five drugs causing most overdose deaths (fentanyl, methamphetamine, cocaine, heroin, and oxycodone).^26^

## Statistical Analysis

Descriptive characteristics were summarized using N (%) for sex and RUCA. Mean (standard deviation) were used for age and SVI. Results with less than 10 observations are reported as <10. Completeness of SDoH data and prevalence of non-canonical values were estimated using logistic regression adjusting for individual age and sex at the data extraction date. The reference racial category was White.

## Results

Table 1. displays the demographic characterization of the 12,696,680 individuals with T2D. This is about a third of all individuals within the U.S. with T2D. The average age was 65.12 (SD: 14.77) years, and 50.1% were female.

**Table 1.**
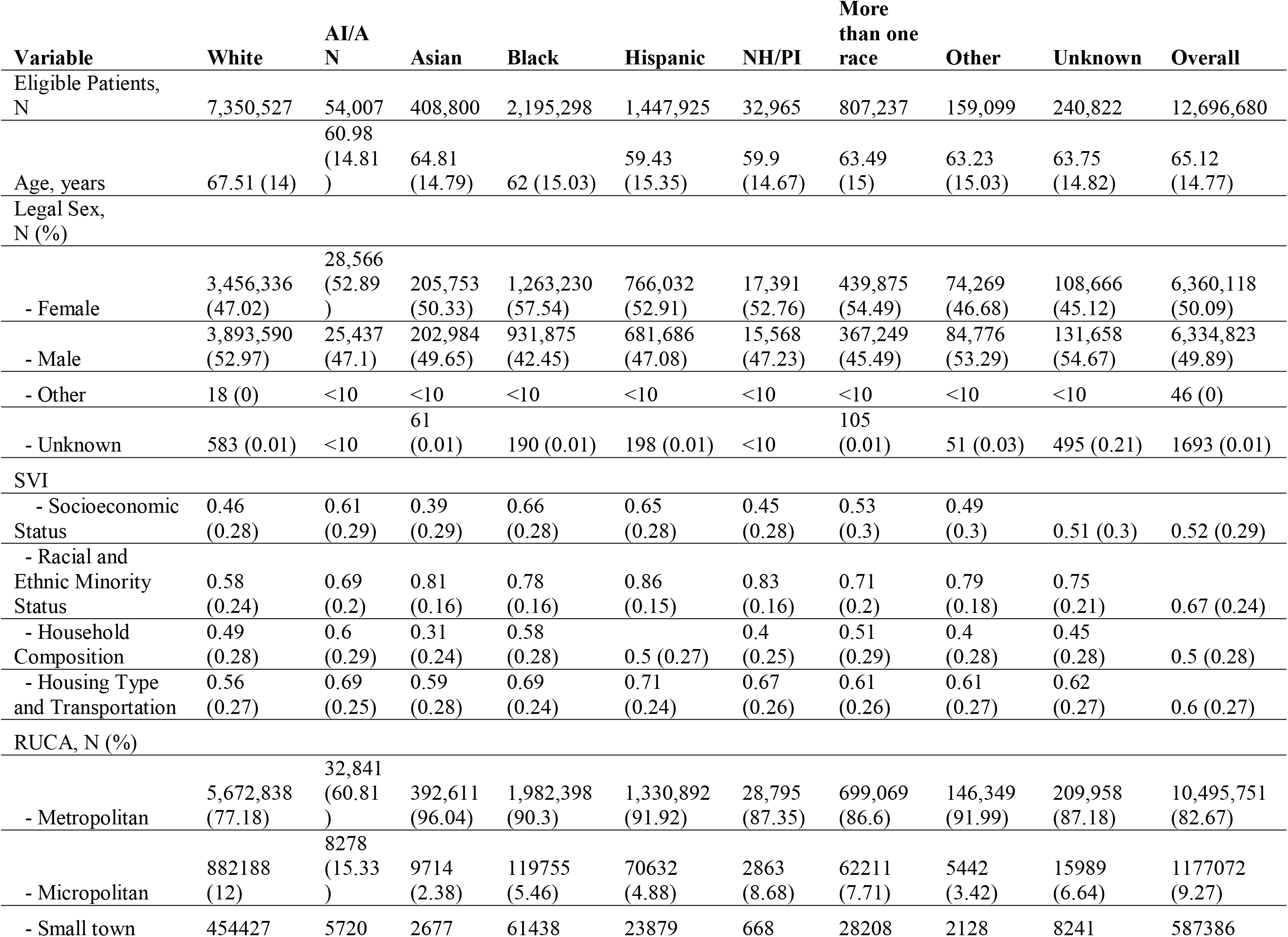
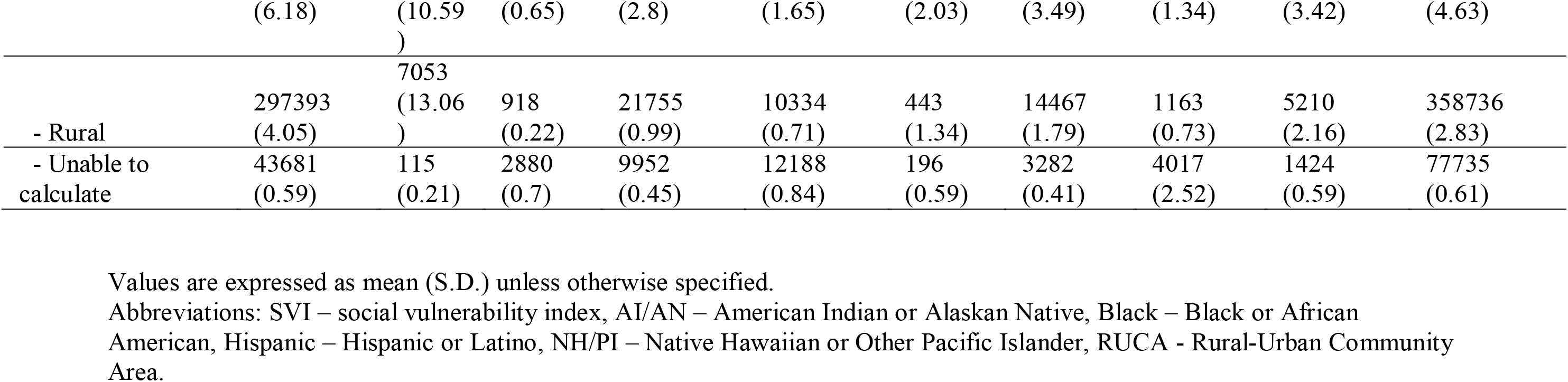
Patient Demographics and Neighborhood-level SDoH by Race/Ethnicity.

Neighborhood-level SDoH (SVI and RUCA) are also summarized in Table 1. SVI was available for 100% of individuals and RUCA was available for 99.4% of individuals.

Completeness of SDoH variables by race/ethnicity is summarized in **Figure 2** and **Supplement Table S1. Figure 3** and **Supplement Table S2** summarize data related to SDoH by race/ethnicity group. Below we discuss the findings for each of the 32 variables evaluated.

**Figure 2.**
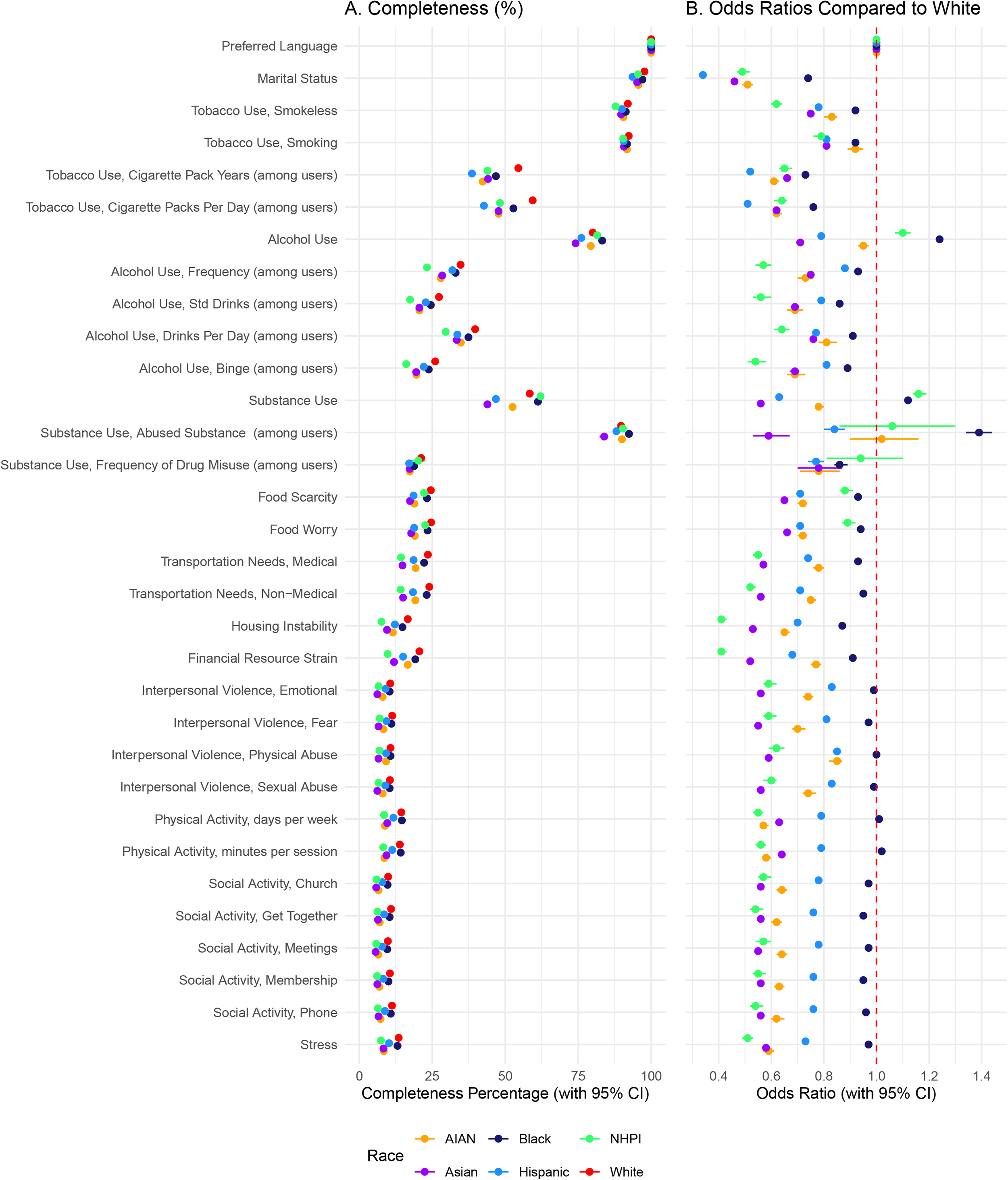
Completeness of SDoH Variables Among Patients with T2D by Race/Ethnicity. A Percentage of complete records. B. Odds ratios compared to White individuals. All values were calculated using logistic regression and adjusted for age and sex. Abbreviations: AI/AN – American Indian or Alaskan Native, Black – Black or African American, Hispanic – Hispanic or Latino, NH/PI – Native Hawaiian or Other Pacific Islander

**Figure 3.**
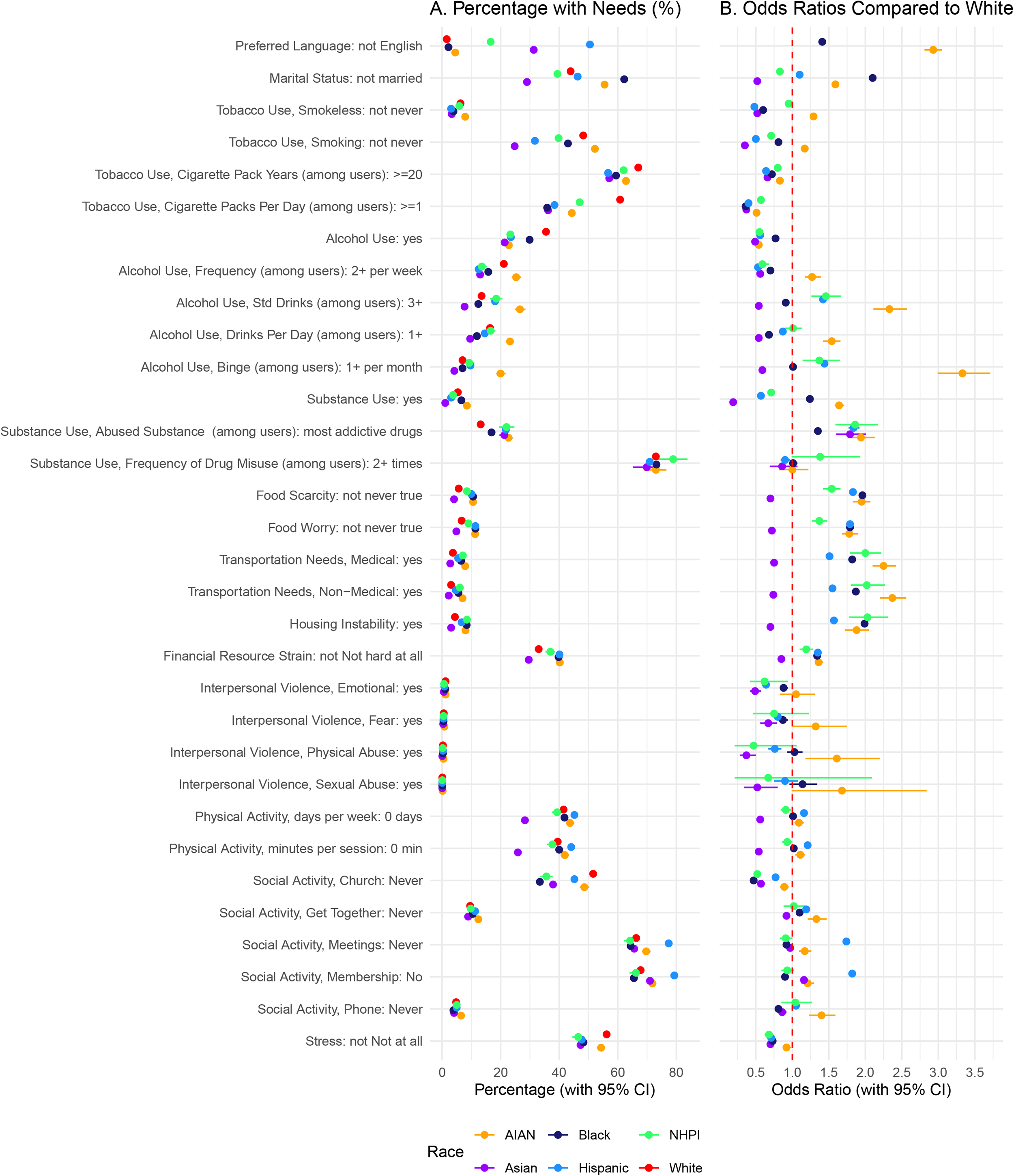
Presence of Issues Among Patients with T2D by Race/Ethnicity. A Percentage of records with issues. B. Odds ratios compared to White individuals. All values were calculated using logistic regression and adjusted for age and sex. Odds ratios above 4 are removed. Abbreviations: AI/AN – American Indian or Alaskan Native, Black – Black or African American, Hispanic – Hispanic or Latino, NH/PI – Native Hawaiian or Other Pacific Islander

### Preferred Language

This variable was universally available, without any race/ethnicity differentiation. The Hispanic group reported a language preference different than the white majority class (English). Asians and NHPIs had a substantial non-White that preferred languages other than English.

### Marital Status

Marital status was reported for 93.6-97.7% of the cohort depending on race/ethnicity. The non-canonical values were reported for between 29.0-62.2% of the racial subgroups. Black (OR: 1.41 [95% CI: 1.39-1.44]) and AI/AN (OR: 1.59 [95% CI: 1.56-1.62]) individuals were more likely not to be married compared to White individuals, while Asian individuals were less likely not to be married (OR: 0.52 [95% CI: 0.52-0.53]).

### Tobacco Use

Data on smoking and smokeless tobacco use were reported for 87.8-92.3% of the subgroups and were available slightly more often for individuals classified as White. Forty-eight percent of T2D individuals reported a positive history of smoking and 6% reported using smokeless tobacco. Among smokers, detailed smoking information was available for over 60% of White group, but less for individuals from racial non-White groups. Fifty five percent of AI/AN with available smoking information reported that they currently smoke of smoked in the past. Asian (OR: 0.52 [95% CI: 0.51-0.53]) and Hispanic (OR: 0.60 [95% CI: 0.60-0.61]) individuals had significantly lower odds of smoking compared to White individuals. Sixty seven percent of White persons had cumulative smoking history of more than 20 pack-years. Among those with non-White race/ethnicity, fewer individuals reported smoking more than 20 pack-years.

### Alcohol Use

Among persons identified as White, completeness on alcohol use was 80.0% [95% CI: 80.0-80.1]. Availability of alcohol exposure was lower for AI/AN (OR: 0.95 [95% CI: 0.93-0.97]), Asian (OR: 0.71 [95% CI: 0.71-0.72]), and Hispanic (OR: 0.79 [95% CI: 0.79-0.8]) individuals and higher for Black (OR: 1.24 [95% CI: 1.23-1.25]) and NH/PI (OR: 1.1 [95% CI: 1.07-1.13]) individuals compared to White individuals (**Table S1**). AI/AN (OR: 0.54 [95% CI: 0.52-0.55]), Asian (OR: 0.49 [95% CI: 0.49-0.50]), Black (OR: 0.77 [95% CI: 0.77-0.78]), Hispanic (OR: 0.56 [95% CI: 0.55-0.56]), and NH/PI (OR: 0.55 [95% CI: 0.53-0.57]) individuals were less likely to consume alcohol compared to White individuals (**Table S2**). Among those who drink, AI/AN reported higher alcohol consumption and more binge drinking.

### Substance Use

Among persons identified as White, the data completeness was 58.4 [95% CI: 58.4-58.5]. Availability of reported substance use was lower for AI/AN, Asian, and Hispanic individuals and higher for Black and NH/PI individuals compared to White individuals. Further, if substance use was reported, Black individuals were more likely to be asked about the type of substance used (OR = 1.35 [95% CI: 1.3-1.39]). Among those with available data, AI/AN (OR: 1.64 [95% CI: 1.58-1.71]) and Black individuals (OR: 1.24 [95% CI: 1.23-1.25]) had higher odds of substance use compared to White individuals, while Asian (OR: 0.19 [95% CI: 0.18-0.2]) and Hispanic (OR: 0.57 [95% CI: 0.56-0.58]) individuals had lower odds. Minority individuals had higher odds of using the five drugs that lead to most overdose deaths.

### Patient needs

Data completeness for food scarcity, food worry, transportation needs, housing instability and financial resource strain was recorded for 7.6% to 24.6% of individuals across race/ethnicity groups. Among those with available data, AI/AN, Black NHPI and Hispanic individuals had higher unmet needs compared to persons identified as White. Asian individuals had lower level of unmet financial needs. AI/AN, Black, Hispanic, and NH/PI individuals experienced higher odds of housing instability and faced greater financial resource strain compared to persons identified as White.

### Interpersonal Violence

Data completeness for interpersonal violence ranged from 6.2% to 11.3%. From those with complete data, less than 1% reported experiencing interpersonal violence. AI/AN individuals reported slightly higher levels of physical abuse. Asian individuals reported slightly lower rates. AI/AN individuals had similar odds of experiencing emotional violence, fear-based violence, and physical abuse compared to White individuals, while Asian, Black, Hispanic, and NH/PI individuals had lower odds of this exposure.

### Physical activity

Data completeness for physical activity was around from 8.2% to 14.4%. Completeness of physical activity data was lower for AI/AN, NH/PI, Asian, and Hispanic individuals. Asian individuals reported high rates of physical activity compared to other groups. More AI/AN and Hispanic individuals reported not exercising. Hispanic and AI/AN individuals were more likely to report 0 minutes of physical activity per session compared to White individuals. In contrast, Asian individuals were less likely to report zero minutes of physical activity.

### Social Activity

Complete data for social activity was available for 5.8% to 11.2% of individuals. All individuals from non-White groups had less complete data than persons identified as White. Black, Asian, Hispanic, and NH/PI individuals were less likely to never attend church compared to White individuals, indicating higher church participation among these groups. Hispanic individuals were more likely to have no meetings, and no memberships documented.

### Stress

Data completeness for stress ranged from 7.4% for NH/PI to 13.5% among White individuals. All individuals of non-White race/ethnicity had less complete data than persons identified as White. Among those with available data, non-White individuals reported less stress than persons identified as White.

## Discussion

This study examines the completeness of Social Determinants of Health (SDoH) data for individuals with T2D in the Epic Cosmos database. Neighborhood-level SDoH variables, such as the SVI and RUCA, are universally available due to the necessity of addresses for administrative purposes. Additionally, individual-level SDoH data—including race/ethnicity, sex, age, preferred language, marital status, and substance use (tobacco, alcohol, and drugs)—are broadly documented. The widespread availability of substance use history is particularly promising, given its significant influence on T2D progression.^27–30^ These data are crucial for identifying individuals eligible for targeted interventions, thereby helping to reduce complications and mortality through treatment, cessation, and screening programs. However, the completeness of substance use data varies by substance type and race/ethnicity, ranging from 43.9% to 92.3%, suggesting that data augmentation methods may be necessary.^31,32^ These findings suggest that equity analyses using SDoH variables are feasible, though caution is warranted.

Conversely, several variables are not yet suitable for AI fairness research due to limited completeness. Data on resource needs, interpersonal violence, physical activity, social connections, and stress are available for only 5.6%-24.6% of individuals, depending on the attribute and race/ethnicity. The low completeness rates for these variables are concerning, as they could provide critical insights into the lived experiences of individuals with T2D, significantly impacting disease management and outcomes. The lack of complete data complicates the understanding of patient challenges and may result in less effective interventions. Significant barriers remain to systematically collecting these data in EHRs. Healthcare teams are already burdened with data collection and entry, and the time required to ask SDoH-related questions and address non-canonical results is substantial. Moreover, reluctance among healthcare teams to inquire about SDoH issues is often due to a lack of resources to address identified problems. Additionally, the frequency of data collection and its impact on outcomes remain unclear.

We observed systemic under-collection of SDoH data in Non-White racial/ethnic groups, which coincided with a higher burden of non-canonical SDoH values compared to White individuals. AI models trained on biased data may inadvertently reinforce existing biases, compromising performance in Non-White groups.^33^ Insufficient and biased SDoH data may also lead to AI systems that fail to accurately represent these groups, perpetuating health disparities and leading to inequities in resource allocation.^33^ Addressing this under-collection is critical for AI fairness, requiring better data collection, augmentation, and continuous bias monitoring. Further research should focus on mitigating varying levels of SDoH data completeness across racial/ethnic groups using statistical and AI methods for data augmentation.^31,32^

SDoH data collection might also be influenced by systemic racism. For instance, Black and Native Hawaiian/Pacific Islander (NH/PI) individuals were more likely to have alcohol and substance use recorded, potentially reflecting differences in data collection practices or clinician biases. Despite higher data completeness for alcohol consumption among Black individuals, reported consumption rates were lower compared to White individuals. This disparity may stem from stereotypes that view Black individuals through a lens of violent behavior and substance abuse. The availability of alcohol and substance use data, coupled with the lack of comprehensive data on other SDoH, may lead to an incomplete understanding of Black and NH/PI individuals’ health needs and less effective interventions.

Our study also identified significant disparities in the prevalence of non-canonical values across different racial and ethnic groups, underscoring the need for targeted interventions to address these inequities. For example, American Indian/Alaska Native (AI/AN) individuals, who experience higher transportation needs, may benefit from programs specifically designed to address and mitigate transportation barriers in these communities. These findings can inform policy decisions and resource allocation, ensuring that interventions and healthcare services are directed toward the populations most in need, ultimately contributing to the development of more effective and sustainable health programs.

## Limitations

This study has several limitations. First, it relies on retrospective EHR data, which are prone to documentation biases and missing information. While the Epic Cosmos database offers a comprehensive source of data from various healthcare environments, the generalizability of our findings may be constrained to individuals treated at institutions using the Epic EHR system. However, given that Epic Cosmos includes data from more than half of the U.S. population, we believe our results are broadly applicable. Second, the definition of SDoH is still evolving, and some of the variables studied (e.g., substance use) might not be included as SDoH by some stakeholders. Third, our study was limited to SDoH variables available in Epic Cosmos. Some essential SDoH variables are not currently collected in EHRs or are unavailable in the Epic Cosmos database. Further, we looked at only SDoH variables recorded in structured, while it is possible to extract SDoH from clinical notes using natural language processing and text mining methods,^34^ notes are not available at Epic Cosmos currently. Fourth, there is a high likelihood that SDoH data are collected differently across healthcare organizations, potentially affecting the validity of the findings. Lastly, we did not use the SUrveillance, PREvention, and ManagEment of Diabetes Mellitus (SUPREME-DM) algorithm, which could have identified more individuals with T2D. However, we focused on individuals with documented T2D diagnoses, using more restrictive inclusion criteria.

## Conclusion

This study highlights the feasibility and importance of systematically collecting SDoH data for individuals with T2D. The widespread availability of neighborhood-level SDoH variables and some individual-level SDoH variables, such as race, ethnicity, legal sex, preferred language, marital status, and substance use, demonstrates the potential for EHR data to support AI equity research. However, the study also identifies concerning gaps in the completeness of critical individual-level SDoH variables such as resource needs, interpersonal violence, physical activity, social connections, and stress. The findings underscore the need to recognize the value of SDoH in healthcare data systems and the importance of equitable data collection.

## Data Availability Statement

Authors have no acknowledgments.

## Data Availability Statement

The data underlying this article cannot be shared publicly due to Epic Cosmos policies.

