## Supplemental Tables for "Characterization and Racial Stratification of Social Determinants of Health for Individuals with Type 2 Diabetes as Recorded in Electronic Health Records: Implications for Artificial Intelligence Development"

### Supplement

**Table S1. Completeness of SDoH Variables Among Patients with T2D by Race/Ethnicity**

| **Variable** | **White** | **AI/AN** | **Asian** | **Black** | **Hispanic** | **NH/PI** |
| --- | --- | --- | --- | --- | --- | --- |
| Preferred Language | 100 | 100, OR: 1 | 100, OR: 1 | 100, OR: 1 | 100, OR: 1 | 100, OR: 1 |
| Marital Status | 97.7 (97.7-97.8) | 95.6 (95.4-95.8), OR: 0.51 (0.49-0.53) | 95.2 (95.1-95.3), OR: 0.46 (0.45-0.47) | 97 (96.9-97), OR: 0.74 (0.73-0.75) | 93.6 (93.5-93.6), OR: 0.34 (0.33-0.34) | 95.5 (95.3-95.7), OR: 0.49 (0.47-0.52) |
| Tobacco Use, Smokeless | 92.0 (92.0-92.1) | 90.5 (90.3-90.8), OR: 0.83 (0.8-0.85) | 89.7 (89.6-89.7), OR: 0.75 (0.74-0.76) | 91.3 (91.3-91.4), OR: 0.92 (0.91-0.92) | 90 (89.9-90), OR: 0.78 (0.77-0.78) | 87.8 (87.4-88.1), OR: 0.62 (0.6-0.64) |
| Tobacco Use, Smoking | 92.3 (92.3-92.3) | 91.7 (91.4-91.9), OR: 0.92 (0.89-0.95) | 90.7 (90.6-90.8), OR: 0.81 (0.81-0.82) | 91.7 (91.6-91.7), OR: 0.92 (0.91-0.92) | 90.6 (90.6-90.7), OR: 0.81 (0.8-0.81) | 90.4 (90.1-90.7), OR: 0.79 (0.76-0.82) |
| - Cigarette Pack Years (among users) | 54.5 (54.4-54.6) | 42.3 (41.7-42.9), OR: 0.61 (0.6-0.63) | 44.1 (43.8-44.4), OR: 0.66 (0.65-0.67) | 46.8 (46.7-46.9), OR: 0.73 (0.73-0.74) | 38.6 (38.4-38.7), OR: 0.52 (0.52-0.53) | 43.9 (43-44.9), OR: 0.65 (0.63-0.68) |
| - Cigarette Packs Per Day (among users) | 59.4 (59.3-59.5) | 47.7 (47.1-48.3), OR: 0.62 (0.61-0.64) | 47.7 (47.3-48), OR: 0.62 (0.61-0.63) | 52.8 (52.7-52.9), OR: 0.76 (0.76-0.77) | 42.7 (42.5-42.8), OR: 0.51 (0.5-0.51) | 48.2 (47.3-49.1), OR: 0.64 (0.61-0.66) |
| Alcohol Use | 80.0 (80.0-80.1) | 79.3 (78.9-79.6), OR: 0.95 (0.93-0.97) | 74.1 (73.9-74.2), OR: 0.71 (0.71-0.72) | 83.2 (83.2-83.3), OR: 1.24 (1.23-1.25) | 76.1 (76-76.2), OR: 0.79 (0.79-0.8) | 81.5 (81.1-82), OR: 1.1 (1.07-1.13) |
| - Drinks Per Day (among users) | 39.7 (39.5-39.8) | 34.8 (33.9-35.7), OR: 0.81 (0.78-0.85) | 33.4 (33-33.7), OR: 0.76 (0.75-0.77) | 37.4 (37.2-37.5), OR: 0.91 (0.9-0.91) | 33.6 (33.4-33.8), OR: 0.77 (0.76-0.78) | 29.6 (28.5-30.7), OR: 0.64 (0.61-0.67) |
| - Frequency (among users) | 34.7 (34.5-34.8) | 27.9 (27-28.7), OR: 0.73 (0.7-0.76) | 28.4 (28.1-28.8), OR: 0.75 (0.73-0.76) | 33 (32.9-33.2), OR: 0.93 (0.92-0.94) | 31.9 (31.8-32.1), OR: 0.88 (0.87-0.89) | 23.2 (22.2-24.2), OR: 0.57 (0.54-0.6) |
| - Standard Drinks (among users) | 27.3 (27.2-27.5) | 20.6 (19.8-21.4), OR: 0.69 (0.66-0.72) | 20.6 (20.3-20.9), OR: 0.69 (0.68-0.7) | 24.5 (24.4-24.6), OR: 0.86 (0.85-0.87) | 22.8 (22.7-23), OR: 0.79 (0.78-0.8) | 17.4 (16.5-18.4), OR: 0.56 (0.53-0.6) |
| - Binge (among users) | 26 (25.9-26.2) | 19.6 (18.8-20.4), OR: 0.69 (0.66-0.73) | 19.5 (19.2-19.8), OR: 0.69 (0.67-0.7) | 23.8 (23.7-23.9), OR: 0.89 (0.88-0.9) | 22.1 (22-22.3), OR: 0.81 (0.8-0.82) | 16.1 (15.2-17), OR: 0.54 (0.51-0.58) |
| Substance Use | 58.4 (58.4-58.5) | 52.5 (52-52.9), OR: 0.78 (0.77-0.8) | 43.9 (43.8-44.1), OR: 0.56 (0.55-0.56) | 61.2 (61.1-61.3), OR: 1.12 (1.12-1.13) | 46.8 (46.8-46.9), OR: 0.63 (0.62-0.63) | 62.1 (61.5-62.6), OR: 1.16 (1.14-1.19) |
| - Abused Substance (among users) | 89.7 (89.5-90) | 90 (88.8-91), OR: 1.02 (0.9-1.16) | 83.9 (82.3-85.3), OR: 0.59 (0.53-0.67) | 92.4 (92.2-92.5), OR: 1.39 (1.34-1.44) | 88.1 (87.7-88.4), OR: 0.84 (0.8-0.88) | 90.3 (88.3-91.9), OR: 1.06 (0.86-1.3) |
| - Frequency of Drug Misuse (among users) | 21.2 (20.8-21.5) | 17.3 (16-18.7), OR: 0.78 (0.71-0.86) | 17.3 (15.9-19), OR: 0.78 (0.7-0.87) | 18.8 (18.6-19.1), OR: 0.86 (0.84-0.89) | 17.1 (16.7-17.5), OR: 0.77 (0.74-0.8) | 20.2 (17.9-22.7), OR: 0.94 (0.81-1.1) |
| Food Scarcity | 24.5 (24.4-24.5) | 18.9 (18.5-19.2), OR: 0.72 (0.7-0.73) | 17.4 (17.3-17.6), OR: 0.65 (0.65-0.66) | 23.2 (23.2-23.3), OR: 0.93 (0.93-0.94) | 18.6 (18.5-18.7), OR: 0.71 (0.7-0.71) | 22.2 (21.8-22.7), OR: 0.88 (0.86-0.91) |
| Food Worry | 24.6 (24.5-24.7) | 19 (18.7-19.3), OR: 0.72 (0.7-0.73) | 17.8 (17.7-17.9), OR: 0.66 (0.66-0.67) | 23.4 (23.3-23.4), OR: 0.94 (0.93-0.94) | 18.8 (18.7-18.9), OR: 0.71 (0.71-0.71) | 22.6 (22.2-23.1), OR: 0.89 (0.87-0.92) |
| Transportation Needs, Medical | 23.5 (23.4-23.5) | 19.3 (19-19.7), OR: 0.78 (0.76-0.8) | 14.8 (14.7-14.9), OR: 0.57 (0.56-0.57) | 22.2 (22.2-22.3), OR: 0.93 (0.93-0.94) | 18.6 (18.5-18.6), OR: 0.74 (0.74-0.75) | 14.3 (14-14.7), OR: 0.55 (0.53-0.56) |
| Transportation Needs, Non-Medical | 24.0 (24.0-24.1) | 19.2 (18.9-19.5), OR: 0.75 (0.74-0.77) | 15.0 (14.9-15.1), OR: 0.56 (0.55-0.56) | 23.1 (23.1-23.2), OR: 0.95 (0.95-0.95) | 18.4 (18.3-18.5), OR: 0.71 (0.71-0.72) | 14.2 (13.8-14.6), OR: 0.52 (0.51-0.54) |
| Housing Instability | 16.6 (16.6-16.7) | 11.5 (11.3-11.8), OR: 0.65 (0.64-0.67) | 9.5 (9.4-9.6), OR: 0.53 (0.52-0.53) | 14.8 (14.7-14.8), OR: 0.87 (0.86-0.87) | 12.2 (12.1-12.3), OR: 0.7 (0.69-0.7) | 7.6 (7.3-7.9), OR: 0.41 (0.4-0.43) |
| Financial Resource Strain | 20.6 (20.5-20.7) | 16.6 (16.3-17), OR: 0.77 (0.75-0.79) | 11.9 (11.8-12), OR: 0.52 (0.51-0.53) | 19.2 (19.1-19.2), OR: 0.91 (0.91-0.92) | 15 (14.9-15), OR: 0.68 (0.67-0.68) | 9.7 (9.3-10), OR: 0.41 (0.4-0.43) |
| Interpersonal Violence, Emotional | 10.6 (10.5-10.6) | 8 (7.8-8.3), OR: 0.74 (0.72-0.76) | 6.2 (6.1-6.3), OR: 0.56 (0.55-0.57) | 10.4 (10.4-10.5), OR: 0.99 (0.98-0.99) | 9 (8.9-9), OR: 0.83 (0.83-0.84) | 6.6 (6.3-6.8), OR: 0.59 (0.57-0.62) |
| Interpersonal Violence, Fear | 11.3 (11.3-11.4) | 8.3 (8-8.5), OR: 0.7 (0.68-0.73) | 6.6 (6.5-6.7), OR: 0.55 (0.54-0.56) | 11 (10.9-11), OR: 0.97 (0.96-0.97) | 9.3 (9.3-9.4), OR: 0.81 (0.8-0.81) | 7 (6.8-7.3), OR: 0.59 (0.57-0.62) |
| Interpersonal Violence, Physical Abuse | 10.7 (10.7-10.8) | 9.2 (9-9.5), OR: 0.85 (0.82-0.87) | 6.6 (6.5-6.7), OR: 0.59 (0.58-0.6) | 10.7 (10.6-10.7), OR: 1 (0.99-1) | 9.2 (9.2-9.3), OR: 0.85 (0.84-0.86) | 6.9 (6.6-7.2), OR: 0.62 (0.59-0.65) |
| Interpersonal Violence, Sexual Abuse | 10.5 (10.5-10.6) | 8.0 (7.8-8.3), OR: 0.74 (0.72-0.77) | 6.2 (6.1-6.3), OR: 0.56 (0.55-0.57) | 10.4 (10.4-10.5), OR: 0.99 (0.98-1) | 8.9 (8.9-9), OR: 0.83 (0.83-0.84) | 6.6 (6.3-6.9), OR: 0.6 (0.57-0.62) |
| Physical activity, days per week | 14.4 (14.4-14.5) | 8.8 (8.6-9.1), OR: 0.57 (0.56-0.59) | 9.6 (9.6-9.7), OR: 0.63 (0.63-0.64) | 14.6 (14.5-14.6), OR: 1.01 (1.01-1.02) | 11.7 (11.7-11.8), OR: 0.79 (0.78-0.79) | 8.5 (8.2-8.8), OR: 0.55 (0.53-0.57) |
| Physical activity, minutes per session | 13.9 (13.8-13.9) | 8.6 (8.4-8.8), OR: 0.58 (0.57-0.6) | 9.3 (9.2-9.4), OR: 0.64 (0.63-0.64) | 14.2 (14.1-14.2), OR: 1.02 (1.02-1.03) | 11.3 (11.3-11.4), OR: 0.79 (0.79-0.8) | 8.2 (8-8.6), OR: 0.56 (0.54-0.58) |
| Social Activity, Church | 9.9 (9.9-10) | 6.6 (6.4-6.8), OR: 0.64 (0.62-0.66) | 5.8 (5.7-5.8), OR: 0.56 (0.55-0.56) | 9.7 (9.6-9.7), OR: 0.97 (0.97-0.98) | 7.9 (7.9-8), OR: 0.78 (0.78-0.79) | 5.9 (5.7-6.2), OR: 0.57 (0.55-0.6) |
| Social Activity, Get Together | 10.9 (10.8-10.9) | 7 (6.8-7.3), OR: 0.62 (0.6-0.64) | 6.4 (6.3-6.4), OR: 0.56 (0.55-0.57) | 10.4 (10.3-10.4), OR: 0.95 (0.94-0.96) | 8.5 (8.5-8.5), OR: 0.76 (0.76-0.77) | 6.2 (6-6.5), OR: 0.54 (0.52-0.57) |
| Social Activity, Meetings | 9.8 (9.8-9.9) | 6.5 (6.3-6.7), OR: 0.64 (0.62-0.66) | 5.6 (5.6-5.7), OR: 0.55 (0.54-0.56) | 9.6 (9.5-9.6), OR: 0.97 (0.96-0.98) | 7.8 (7.8-7.9), OR: 0.78 (0.77-0.79) | 5.8 (5.6-6.1), OR: 0.57 (0.54-0.6) |
| Social Activity, Membership | 10.5 (10.5-10.6) | 6.9 (6.7-7.1), OR: 0.63 (0.61-0.65) | 6.2 (6.1-6.3), OR: 0.56 (0.55-0.57) | 10 (10-10), OR: 0.95 (0.94-0.95) | 8.2 (8.2-8.3), OR: 0.76 (0.76-0.77) | 6.1 (5.8-6.4), OR: 0.55 (0.53-0.58) |
| Social Activity, Phone | 11.2 (11.1-11.2) | 7.3 (7.1-7.5), OR: 0.62 (0.6-0.65) | 6.6 (6.5-6.7), OR: 0.56 (0.56-0.57) | 10.8 (10.7-10.8), OR: 0.96 (0.95-0.97) | 8.8 (8.7-8.8), OR: 0.76 (0.76-0.77) | 6.4 (6.1-6.6), OR: 0.54 (0.52-0.57) |
| Stress | 13.5 (13.4-13.5) | 8.4 (8.2-8.7), OR: 0.59 (0.57-0.61) | 8.3 (8.2-8.4), OR: 0.58 (0.57-0.59) | 13.1 (13.1-13.2), OR: 0.97 (0.96-0.98) | 10.2 (10.1-10.2), OR: 0.73 (0.72-0.73) | 7.4 (7.1-7.7), OR: 0.51 (0.49-0.53) |

Values for White are expressed as estimated mean with 95% confidence intervals (CIs). Values for AI/AN, Asian, Black, Hispanic, and NW/PI are reported as estimated mean with 95% CI, followed by odds ratios (ORs) with 95% confidence intervals (CI). All values were calculated using logistic regression and adjusted for age and sex.

Abbreviations: AI/AN – American Indian or Alaskan Native, Black – Black or African American, Hispanic – Hispanic or Latino, NH/PI – Native Hawaiian or Other Pacific Islander

**Table S2. Percentage of Non-canonical Values Among Patients with T2D by Race/Ethnicity**

| **Variable** | **Non-Canonical Values** | **White** | **AI/AN** | **Asian** | **Black** | **Hispanic** | **NH/PI** |
| --- | --- | --- | --- | --- | --- | --- | --- |
| Preferred Language | Not “English” | 1.6 (1.6-1.6) | 4.5 (4.3-4.7), OR: 2.93 (2.81-3.05) | 31.3 (31.2-31.5), OR: 28.37 (27.98-28.77) | 2.2 (2.2-2.2), OR: 1.41 (1.39-1.44) | 50.5 (50.4-50.6), OR: 63.42 (62.61-64.23) | 16.6 (16.2-17), OR: 12.34 (11.95-12.74) |
| Marital Status | Not “Married” | 43.9 (43.8-44) | 55.5 (55.1-55.9), OR: 1.59 (1.56-1.62) | 29 (28.9-29.2), OR: 0.52 (0.52-0.53) | 62.2 (62.2-62.3), OR: 2.1 (2.1-2.11) | 46.3 (46.3-46.4), OR: 1.1 (1.1-1.11) | 39.4 (38.9-40), OR: 0.83 (0.81-0.85) |
| Tobacco Use, Smokeless | Not “Never” | 6.3 (6.2-6.3) | 7.9 (7.7-8.2), OR: 1.29 (1.25-1.34) | 3.3 (3.3-3.4), OR: 0.52 (0.51-0.53) | 3.9 (3.8-3.9), OR: 0.6 (0.6-0.61) | 3.1 (3.1-3.2), OR: 0.48 (0.48-0.49) | 5.9 (5.7-6.2), OR: 0.95 (0.9-0.99) |
| Tobacco Use, Smoking | Not “Never” | 48.2 (48.2-48.3) | 52.2 (51.7-52.6), OR: 1.17 (1.15-1.19) | 24.8 (24.6-24.9), OR: 0.35 (0.35-0.36) | 43 (42.9-43), OR: 0.81 (0.8-0.81) | 31.7 (31.6-31.8), OR: 0.5 (0.5-0.5) | 39.8 (39.2-40.3), OR: 0.71 (0.69-0.72) |
| - Cigarette Pack Years (among users) | >=20 | 67.0 (66.9-67.2) | 62.8 (61.9-63.7), OR: 0.83 (0.8-0.86) | 57.1 (56.7-57.6), OR: 0.66 (0.64-0.67) | 59.4 (59.3-59.6), OR: 0.72 (0.71-0.73) | 56.7 (56.5-57), OR: 0.64 (0.64-0.65) | 62 (60.7-63.3), OR: 0.8 (0.76-0.85) |
| - Cigarette Packs Per Day (among users) | >=1 | 60.8 (60.7-61) | 44.3 (43.4-45.2), OR: 0.51 (0.49-0.53) | 36.2 (35.8-36.7), OR: 0.37 (0.36-0.37) | 35.9 (35.8-36.1), OR: 0.36 (0.36-0.36) | 38.4 (38.2-38.7), OR: 0.4 (0.4-0.41) | 47 (45.7-48.3), OR: 0.57 (0.54-0.6) |
| Alcohol Use: | Not “Never used” | 35.5 (35.5-35.6) | 22.8 (22.4-23.2), OR: 0.54 (0.52-0.55) | 21.4 (21.2-21.5), OR: 0.49 (0.49-0.50) | 29.9 (29.8-29.9), OR: 0.77 (0.77-0.78) | 23.5 (23.4-23.6), OR: 0.56 (0.55-0.56) | 23.3 (22.8-23.8), OR: 0.55 (0.53-0.57) |
| - Drinks Per Day (among users) | >=1 per day | 16.4 (16.2-16.6) | 23.2 (21.9-24.6), OR: 1.54 (1.42-1.66) | 9.6 (9.2-9.9), OR: 0.54 (0.52-0.56) | 11.9 (11.7-12), OR: 0.68 (0.67-0.7) | 14.6 (14.4-14.8), OR: 0.87 (0.85-0.89) | 16.6 (15.1-18.2), OR: 1.01 (0.9-1.13) |
| - Frequency (among users) | >=2 per week | 21.1 (20.8-21.3) | 25.3 (23.8-27), OR: 1.27 (1.17-1.39) | 13 (12.6-13.5), OR: 0.56 (0.54-0.59) | 15.8 (15.6-15.9), OR: 0.7 (0.69-0.71) | 12.5 (12.2-12.7), OR: 0.53 (0.52-0.55) | 13.6 (12.1-15.4), OR: 0.59 (0.51-0.68) |
| - Standard Drinks (among users) | >=3 | 13.5 (13.2-13.7) | 26.6 (24.8-28.5), OR: 2.33 (2.11-2.57) | 7.7 (7.3-8.1), OR: 0.54 (0.51-0.57) | 12.4 (12.2-12.6), OR: 0.91 (0.89-0.93) | 18.1 (17.8-18.4), OR: 1.42 (1.39-1.46) | 18.5 (16.4-20.6), OR: 1.46 (1.26-1.67) |
| - Binge (among users) | >=1 per month | 7 (6.8-7.1) | 20 (18.4-21.7), OR: 3.33 (2.99-3.71) | 4.2 (3.9-4.5), OR: 0.59 (0.54-0.63) | 7 (6.9-7.2), OR: 1.01 (0.98-1.04) | 9.7 (9.5-10), OR: 1.44 (1.39-1.48) | 9.3 (7.9-11), OR: 1.37 (1.14-1.65) |
| Substance Use | Not “Never used” | 5.4 (5.3-5.4) | 8.5 (8.2-8.8), OR: 1.64 (1.58-1.71) | 1.1 (1-1.1), OR: 0.19 (0.18-0.2) | 6.6 (6.5-6.6), OR: 1.24 (1.23-1.25) | 3.1 (3.1-3.1), OR: 0.57 (0.56-0.58) | 3.8 (3.6-4.1), OR: 0.71 (0.66-0.75) |
| - Abused Substance (among users) | fentanyl, methamphetamine, cocaine, heroin, or oxycodone | 13.2 (12.8-13.5) | 22.7 (21.1-24.3), OR: 1.94 (1.76-2.13) | 21.3 (19.6-23.2), OR: 1.79 (1.6-2.01) | 16.9 (16.7-17.2), OR: 1.35 (1.3-1.39) | 21.8 (21.3-22.3), OR: 1.84 (1.77-1.92) | 22 (19.5-24.7), OR: 1.86 (1.59-2.17) |
| - Frequency of Drug Misuse (among users) | >=2 | 73 (72.1-73.9) | 73 (69-76.6), OR: 1 (0.82-1.22) | 69.9 (65.2-74.2), OR: 0.86 (0.69-1.07) | 73.2 (72.6-73.8), OR: 1.01 (0.96-1.07) | 70.9 (69.6-72.1), OR: 0.9 (0.84-0.97) | 78.9 (72.9-83.8), OR: 1.38 (0.99-1.93) |
| Food Scarcity | Not “Never true” | 5.7 (5.7-5.8) | 10.6 (10-11.2), OR: 1.95 (1.83-2.07) | 4.1 (3.9-4.2), OR: 0.7 (0.67-0.73) | 10.6 (10.5-10.7), OR: 1.96 (1.92-1.99) | 10 (9.9-10.1), OR: 1.83 (1.8-1.87) | 8.5 (8-9.2), OR: 1.54 (1.42-1.66) |
| Food Worry | Not “Never true” | 6.7 (6.6-6.8) | 11.3 (10.8-12), OR: 1.78 (1.68-1.9) | 4.9 (4.8-5.1), OR: 0.72 (0.7-0.75) | 11.4 (11.3-11.5), OR: 1.79 (1.76-1.82) | 11.4 (11.3-11.5), OR: 1.79 (1.76-1.83) | 9 (8.4-9.6), OR: 1.37 (1.27-1.48) |
| Transportation Needs, Medical | Not “No issues” | 3.7 (3.6-3.7) | 7.9 (7.4-8.4), OR: 2.25 (2.1-2.42) | 2.8 (2.7-2.9), OR: 0.75 (0.71-0.79) | 6.5 (6.4-6.6), OR: 1.82 (1.78-1.86) | 5.5 (5.4-5.5), OR: 1.51 (1.47-1.55) | 7.1 (6.4-7.8), OR: 2 (1.79-2.22) |
| Transportation Needs, Non-Medical | Not “No issues” | 3.1 (3-3.2) | 7 (6.6-7.5), OR: 2.37 (2.2-2.56) | 2.3 (2.2-2.4), OR: 0.74 (0.7-0.78) | 5.6 (5.6-5.7), OR: 1.87 (1.83-1.91) | 4.7 (4.6-4.8), OR: 1.55 (1.51-1.59) | 6.1 (5.4-6.7), OR: 2.02 (1.8-2.27) |
| Housing Instability | Not “No issues” | 4.4 (4.3-4.5) | 8 (7.4-8.6), OR: 1.88 (1.72-2.05) | 3.1 (2.9-3.3), OR: 0.7 (0.66-0.74) | 8.4 (8.3-8.5), OR: 1.99 (1.94-2.03) | 6.7 (6.6-6.8), OR: 1.57 (1.53-1.61) | 8.5 (7.6-9.6), OR: 2.03 (1.78-2.31) |
| Financial Resource Strain | Not “Not hard at all” | 33 (32.9-33.2) | 40.2 (39.2-41.2), OR: 1.36 (1.31-1.42) | 29.6 (29.2-30), OR: 0.85 (0.83-0.87) | 39.8 (39.7-40), OR: 1.34 (1.33-1.36) | 40.1 (39.8-40.3), OR: 1.35 (1.34-1.37) | 37 (35.3-38.6), OR: 1.19 (1.1-1.28) |
| Interpersonal Violence, Emotional | Not “No issues” | 1.2 (1.1-1.2) | 1.2 (1-1.6), OR: 1.05 (0.83-1.31) | 0.6 (0.5-0.7), OR: 0.49 (0.42-0.57) | 1.1 (1-1.1), OR: 0.88 (0.84-0.94) | 0.8 (0.7-0.8), OR: 0.64 (0.59-0.68) | 0.7 (0.5-1.1), OR: 0.62 (0.42-0.94) |
| Interpersonal Violence, Fear | Not “No issues” | 0.6 (0.6-0.6) | 0.8 (0.6-1), OR: 1.32 (1-1.75) | 0.4 (0.3-0.5), OR: 0.67 (0.56-0.79) | 0.5 (0.5-0.6), OR: 0.87 (0.81-0.94) | 0.5 (0.4-0.5), OR: 0.8 (0.73-0.87) | 0.5 (0.3-0.7), OR: 0.75 (0.46-1.23) |
| Interpersonal Violence, Physical Abuse | Not “No issues” | 0.3 (0.3-0.4) | 0.5 (0.4-0.7), OR: 1.61 (1.18-2.2) | 0.1 (0.1-0.2), OR: 0.37 (0.28-0.5) | 0.3 (0.3-0.4), OR: 1.03 (0.93-1.14) | 0.3 (0.2-0.3), OR: 0.76 (0.67-0.85) | 0.2 (0.1-0.3), OR: 0.47 (0.21-1.06) |
| Interpersonal Violence, Sexual Abuse | Not “No issues” | 0.1 (0.1-0.1) | 0.2 (0.1-0.3), OR: 1.68 (0.99-2.84) | 0.1 (0-0.1), OR: 0.52 (0.34-0.8) | 0.1 (0.1-0.1), OR: 1.14 (0.96-1.34) | 0.1 (0.1-0.1), OR: 0.9 (0.75-1.08) | 0.1 (0-0.2), OR: 0.67 (0.21-2.09) |
| Physical activity, days per week | 0 days | 41.5 (41.3-41.7) | 43.7 (42.3-45.2), OR: 1.09 (1.03-1.16) | 28.3 (27.9-28.8), OR: 0.56 (0.54-0.57) | 41.8 (41.6-42), OR: 1.01 (1-1.02) | 45.2 (44.9-45.4), OR: 1.16 (1.15-1.18) | 39.3 (37.5-41.1), OR: 0.91 (0.84-0.98) |
| Physical activity, minutes per session | 0 minutes | 39.5 (39.2-39.7) | 41.9 (40.5-43.3), OR: 1.11 (1.04-1.17) | 25.9 (25.4-26.3), OR: 0.54 (0.52-0.55) | 40 (39.9-40.2), OR: 1.02 (1.01-1.04) | 44.1 (43.9-44.3), OR: 1.21 (1.19-1.23) | 37.7 (35.8-39.5), OR: 0.93 (0.86-1) |
| Social Activity, Church | Not never | 51.6 (51.3-51.8) | 48.6 (47-50.3), OR: 0.89 (0.83-0.95) | 37.9 (37.3-38.5), OR: 0.57 (0.56-0.59) | 33.4 (33.2-33.6), OR: 0.47 (0.46-0.48) | 45.2 (44.9-45.5), OR: 0.77 (0.76-0.79) | 35.6 (33.5-37.8), OR: 0.52 (0.47-0.57) |
| Social Activity, Get Together | “Never” | 9.6 (9.5-9.8) | 12.4 (11.4-13.5), OR: 1.33 (1.21-1.47) | 8.9 (8.6-9.3), OR: 0.92 (0.88-0.97) | 10.5 (10.3-10.6), OR: 1.1 (1.07-1.12) | 11.3 (11.1-11.4), OR: 1.19 (1.17-1.22) | 9.8 (8.6-11.1), OR: 1.02 (0.88-1.18) |
| Social Activity, Meetings | “Never” | 66.3 (66.1-66.6) | 69.7 (68.2-71.2), OR: 1.17 (1.09-1.26) | 65.6 (65-66.2), OR: 0.97 (0.94-1) | 64.4 (64.1-64.6), OR: 0.92 (0.9-0.93) | 77.4 (77.1-77.6), OR: 1.74 (1.7-1.77) | 64.2 (62.1-66.4), OR: 0.91 (0.83-1) |
| Social Activity, Membership | “No” | 67.8 (67.6-68) | 71.8 (70.3-73.2), OR: 1.21 (1.12-1.3) | 71 (70.5-71.6), OR: 1.16 (1.13-1.2) | 65.5 (65.3-65.7), OR: 0.9 (0.89-0.91) | 79.3 (79.1-79.5), OR: 1.82 (1.79-1.85) | 66.2 (64-68.2), OR: 0.93 (0.85-1.02) |
| Social Activity, Phone | “Never” | 4.8 (4.7-4.9) | 6.5 (5.8-7.4), OR: 1.4 (1.23-1.59) | 4.1 (3.9-4.4), OR: 0.86 (0.81-0.92) | 3.9 (3.8-4), OR: 0.81 (0.78-0.83) | 5 (4.9-5.1), OR: 1.05 (1.02-1.09) | 5 (4.1-6), OR: 1.04 (0.85-1.27) |
| Stress | Not “Not at all stressed” | 56.2 (56-56.4) | 54.3 (52.8-55.7), OR: 0.92 (0.87-0.98) | 47.3 (46.8-47.9), OR: 0.7 (0.68-0.72) | 48.3 (48.1-48.5), OR: 0.73 (0.72-0.74) | 47.7 (47.5-48), OR: 0.71 (0.7-0.72) | 46.5 (44.5-48.5), OR: 0.68 (0.62-0.73) |

Values for White are expressed as estimated mean with 95% confidence intervals (C.I.s). Values for AI/AN, Asian, Black, Hispanic, and NW/PI are reported as estimated mean with 95% CI, followed by odds ratios (ORs) with 95% confidence intervals (CIs). All values were calculated using logistic regression and adjusted for age and sex.

Abbreviations: AI/AN – American Indian or Alaskan Native, Black – Black or African American, Hispanic – Hispanic or Latino, NH/PI – Native Hawaiian or Other Pacific Islander
